# Should Household Air Quality Monitoring Be Considered in Selected Asthma & COPD Patients?

**DOI:** 10.1101/2025.09.11.25335550

**Authors:** Rubèn González-Colom, Alba Gómez-López, Alicia Aguado, Néstor Soler, Núria Sánchez-Ruano, Marta Sorribes, Antonio Montilla-Ibarra, Maria Figols, Alberto Rodríguez, Emili Vela, Jordi Piera-Jiménez, Ramon Farré, Josep Roca, Isaac Cano, Jose Fermoso, Ebymar Arismendi

## Abstract

**Background:** Indoor air quality (IAQ) is a well demonstrated actionable determinant of health status. Low-cost sensors (LCS) could enable patient-centred assessments, but real-world clinical utility is uncertain.

**Objective:** Evaluate the feasibility, usability, and clinical indications of home IAQ monitoring with LCS.

**Methods:** We conducted a cohort study involving household continuous IAQ monitoring with LCS of 205 adults with COPD, bronchiectasis, or asthma. Household IAQ was profiled prospectively over two-month period registering concentrations of CO_2_, PM_2.5_and formaldehyde every 10 min. For each pollutant, dwellings were classified as Good, Moderate or Unhealthy according to the Global Open Air Quality Standards thresholds (GO-AQS). Pulmonary exacerbations requiring unplanned hospitalizations, and all-cause emergency department (ED) visits, over the preceding 12 months were registered and potential relationships with household IAQ results were explored.

**Results:** More than half of homes (51.7%) had at least one pollutant in an at-risk category. The burden was mostly generated by PM_2.5_: 40.1% of dwellings were classified as at risk (32.8% Moderate; 7.3% Unhealthy). Formaldehyde exceeded the low-risk threshold in 22 homes (12.4%). Tobacco smoking, either active or passive, was significantly associated with PM_2.5_levels (p<0.001). No relationships were found between IAQ categories and hospitalizations nor with all-cause ED visits.

**Conclusions:** LCS are useful tools for short-term, targeted household IAQ screening in chronic respiratory patients. Indoor pollution is highly prevalent and largely PM_2.5_driven. Further research is needed to assess the short-term health impacts of these exposures.

**Registration:** NCT06421402.

**What is already known on this topic:** Indoor air pollution constitutes a major environmental determinant of respiratory morbidity, with evidence indicating that reducing exposure through improved IAQ can attenuate associated health risks. The potential of LCS for identifying modifiable exposures in patients with chronic respiratory disease has been suggested, but their real-world feasibility and clinical value to inform preventive care interventions are still uncertain.

**What this study adds:** This study demonstrates that LCS are feasible and reliable tools for short-term household IAQ screening in chronic respiratory patients. Whereas, indoor pollution was found to be highly prevalent, mainly driven by fine PM and indoor smoking, supporting their use for targeted risk assessment.

**How this study might affect research, practice, or policy:** These findings highlight the need to consider IAQ as a modifiable component of chronic respiratory care and support the inclusion of environmental assessments in preventive management. Further research should explore short-term health impacts and develop actionable guidelines for integrating IAQ monitoring into clinical and public health strategies.

## Introduction

Exposure to air pollution is considered the most important environmental health risk factor with significant deleterious impacts on physiological developments during early life and accelerated functional decline in the elderly^1^. There is also robust evidence of both acute and sustained adverse effects of air pollution on several organs and systems, with respiratory disorders being one of the primary concerns^2–6^. Specifically, the effects of exposure to particulate matter (PM), as well as oxidants like nitrogen oxides (NOx) and ozone (O_3_), have been extensively demonstrated^7^. The exposure to formaldehyde (CH_2_O) is also associated with asthma diagnosis and exacerbations^8,9^. Furthermore, there is growing attention on the negative impact of other airborne pollutants, such as volatile organic compounds (VOCs)^10^.

Due to the magnitude of the burden of air pollution on human health, international agencies^11–14^are actively deploying public health policy actions. In this context, research on indoor air quality (IAQ) is raising interest due to unknowns on the sources of indoor pollution^15,16^, the interactions between IAQ and outdoor air quality (OAQ), and most importantly, the lack of available information to generate appropriate regulations and health policies on IAQ, as reported in two recent official statements of the American Thoracic Society (ATS)^17,18^. Likewise, the European Union launched the IDEAL Cluster in 2022^19^, an ambitious research and innovation program encompassing seven consortia working on coordinated action plans to set IAQ standards. One of the relevant areas of action of the IDEAL cluster is to explore the potential of low-cost sensors (LCS) for IAQ monitoring in health-related applications. LCS are gaining traction due to their potential for remote continuous IAQ monitoring of different pollutants, affordability, applicability, and potential for widespread deployment in different scenarios^20–22^.The harmful effects of IAQ on respiratory health have been consistently proven in patients with chronic obstructive pulmonary disease (COPD)^5,23–25^showing significant associations between indoor pollutants and symptoms, functional capacity and risk of exacerbations.

Interestingly, the CLEAN AIR^23^study showed potential health benefits of portable particulate air cleaners. A recent report highlighted that the harmful effects of PM pollution on cardiovascular health in patients with COPD can be mitigated by reducing exposure^4^, suggesting a potential role for household IAQ monitoring in high-risk patients. As stated, LCS might open a window of opportunity to enhance the management of selected patients with chronic obstructive respiratory diseases. It is acknowledged, however, that there are uncertainties regarding the quality of LCS measurements, and their potential for applicability in healthcare^20–22^. Accordingly, our primary objective was to evaluate the feasibility, and usability, of home IAQ monitoring with LCS, as well as to explore their usefulness for clinical management^26–28^.

## Method

### Study cohort

Between October 2023 and March 2025, we enrolled 205 patients with COPD or bronchiectasis or asthma from two complementary settings. Most participants (n = 152; 77%) were recruited through four primary care centres in the Barcelona-Esquerra Integrated Care Area (AISBE)^29^, each serving approximately 20,000 residents. These patients were identified in the Catalan Health Surveillance System (CHSS)^30^with a diagnosis ICD-10-CM^31^J44 (COPD), J45 (asthma) and J47 (bronchiectasis) and classified as high-risk due to disease severity and/or multimorbidity, defined as an Adjusted Morbidity Groups (AMG)^32–34^score at or above the 80th percentile of the regional risk pyramid. The remaining patients (n=53; 23%), all with stage 5 or 6 asthma^35,36^, were enrolled at the Severe Asthma Unit at Hospital Clinic de Barcelona.

Eligibility criteria, recruitment procedures, and monitoring protocols are detailed in the published study protocol^28^. Throughout the study, clinical management followed international recommendations for COPD^37,38^, bronchiectasis^39^, asthma^35,36^management.

### Study design and IAQ evaluation

Conducted and reported in accordance with STROBE^40^guidelines. The study design consisted of the prospective characterization of household IAQ in the study cohort using continuous LCS measurements during a two-month period. The relationships between dwelling IAQ levels and unplanned hospitalizations due to respiratory exacerbations, as well as all-cause emergency department visits, occurred during the previous twelve months were explored (**Figure 1**).

**Figure 1:**
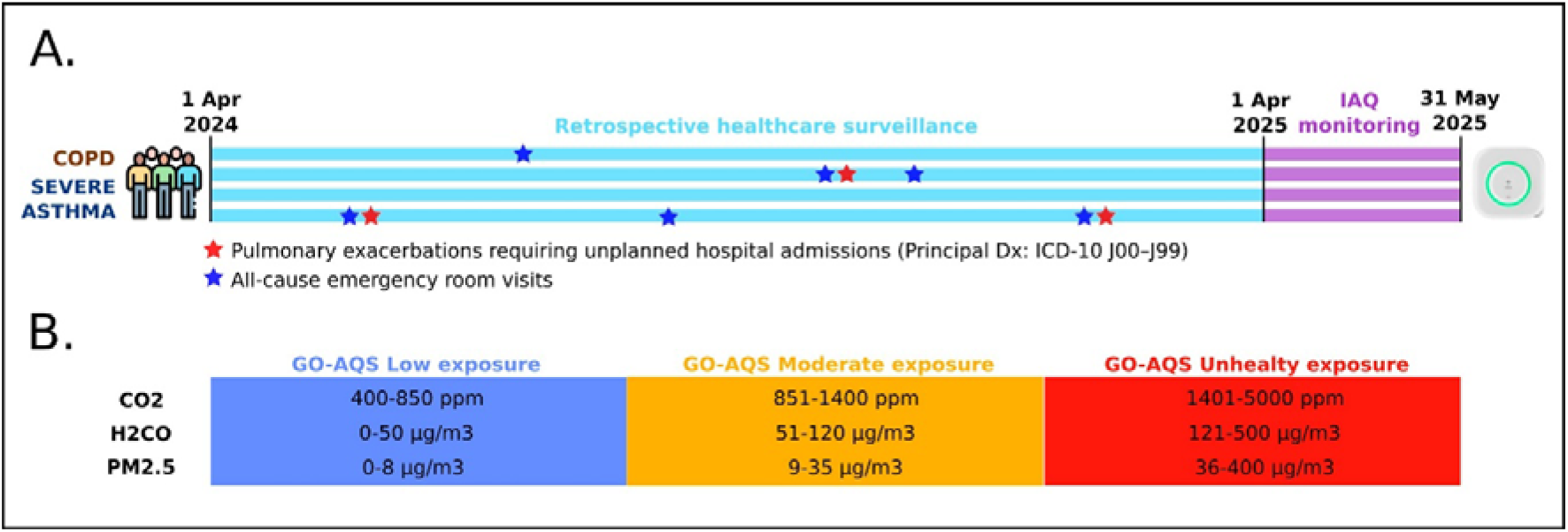
Study timeline and design. (A) Prospective in-home IAQ monitoring (purple) was conducted from 1 April to 31 May 2025. Retrospective surveillance of acute respiratory events (light blue; 1 April 2024–31 March 2025): severe pulmonary exacerbations leading to unplanned hospital admissions (ICD-10-CM J00–J99; red stars) and all-cause emergency department visits (blue stars). (B) Long-term exposure thresholds from the Global Open Air Quality Standards (GO-AQS) used to classify household IAQ as Good, Moderate, or Unhealthy for CO_2_, formaldehyde, and PM_2.5_.

#### Prospective in-home IAQ monitoring (1 Apr 2025 – 31 May 2025)

Between 1 April and 31 May 2025, prospective in-home IAQ monitoring was conducted in patient dwellings using MICA-IBIOT’s^41^LCS to continuously record key IAQ parameters every 10 minutes, including temperature, relative humidity, CO_2_, PM1, PM2.5, PM10, formaldehyde and total VOCs concentrations. The **Supplementary Material**contents detailed information on the manufacturer specifications of the used IAQ sensors and details the validation work conducted within the K-HEALTHinAIR project^42^, comprising chamber tests and in-field comparisons, to assess accuracy, linearity, inter-sensor agreement, and drift. In short, the sensor performance for PM and formaldehyde met predefined quality criteria and was adequate for the study objectives; by contrast, total VOC measurements failed quality thresholds and were excluded from analysis.

To assess indoor pollution levels and stratify homes according to their IAQ quality for different parameters, we applied the reference framework outlined in the Global Open Air Quality Standards (GO AQS) white paper^43^. Subsequently, dwellings were classified using the GO-AQS framework adapted for long-term exposure assessment of the key pollutants monitored: CO_2_, formaldehyde, and PM2.5. The concentration of each pollutant in the patient’s home were classified in three categories: i) Good, ii) Moderate, or iii) Unhealthy ranges. Homes were classified based on their all-time average pollutant concentrations and stratified according to their IAQ status for each parameter. Additionally, the relative frequency of time spent in each risk category was calculated to provide a more detailed analysis of exposure patterns and variability^44,45^. For IAQ analyses, we excluded the cases with less than 20 complete monitoring days and those patients who discontinued follow-up.

#### Retrospective clinical surveillance (1 Apr 2024 – 31 Mar 2025)

To examine the associations between household indoor pollution and severe health events, we retrieved from the CHSS the registries of acute care events over the preceding year (1 Apr 2024 – 31 Mar 2025). The primary outcome was severe pulmonary exacerbations requiring unplanned hospital admission with a principal respiratory diagnosis (ICD-10-CM J00–J99). Planned admissions and non-respiratory urgent admissions were excluded. All-cause emergency department visits were captured as an exploratory outcome.

### Statistical analysis

Numeric variables are described as mean and standard deviation (SD) or median and interquartile range (IQR, Q1–Q3), depending on their distribution. Categorical variables are summarized as counts and percentages; n (%). The comparison of numerical outcomes between patients with respiratory disease exacerbations requiring hospital admission and those who did not was conducted using Student’s t-test for normally distributed variables or the Mann-Whitney U test for non-normally distributed variables. The frequency distributions of categorical factors were compared using Fisher’s exact test. A p-value of <0.05 was considered statistically significant. All statistical analyses were performed using R, version 4.1.1^46^.

### Ethics approval and consent to participate

The Ethical Committee for Human Research at the Hospital Clínic de Barcelona approved the core study protocol for K-HEALTHinAIR on June 29, 2023 (HCB/2023/0126). The study design aligns with the data minimisation principles, ensuring that only the data strictly necessary for the research are collected and utilised. It will be carried out per the Declaration of Helsinki and adheres to the protocol and applicable legal requirements, including the Biomedical Research Act 14/2007 of July 3.

All participants in the study had to sign an informed consent form before undergoing any procedures. They were informed that they could withdraw their consent at any point, without affecting their relationship with their physician or compromising their medical treatment. The study has been registered at ClinicalTrials.gov(Identifier: NCT06421402).

## Results

### Characteristics of the study cohort

During the prospective IAQ monitoring window reported in this manuscript, 27 participants out of the 205 patients included in the cohort (13.2%) were not evaluable, yielding a final analytic sample of 178 patients (86.8%). Of these, 130 (73.0%) were from the community program and 48 (27.0%) from the Severe Asthma Unit. The exclusion reasons are the following: 19 (9.3%) dropped out, 3 (1.5%) died, and 5 (2.4%) were excluded because of IAQ sensor malfunction or insufficient data.

Table 1. summarises baseline characteristics of the study cohort (n=178), stratified by patients with one or more unplanned hospitalisation due to a severe pulmonary exacerbation (n=50; 28.1%) versus those without such events (n=128; 71.9%). Over the one-year retrospective surveillance, 69/178 (38.8%) patients accounted for 113 hospitalisations: 31/113 (27.4%) were planned and 82/113 (72.6%) were urgent. Among urgent admissions, 68/82 (82.9%) were due to respiratory exacerbations, corresponding to 50/178 (28.1%) patients; the remaining 14/82 (17.1%) urgent admissions were for non-respiratory causes.

As displayed in **Table 1**, the patients who experienced at least one unplanned respiratory hospitalisation due to a pulmonary exacerbation along the previous year before the IAQ assessment (n=50) had a substantially higher multimorbidity burden (AMG scoring) and more severe airflow limitation than those without events (n=128). The mean AMG score was higher in the hospitalised group (p<.001), with an over-representation in the very-high-risk band (p=.007). Prior healthcare use in the preceding year was markedly higher among patients with subsequent unplanned admissions: All-cause hospitalisations (p<.001), unplanned hospitalisation (p<.001), and greater total healthcare expenditure (p<.001). Spirometry showed worse obstruction among hospitalised patients: lower FEV_1_z-score (p=.009) and lower FEV_1_/FVC (p<.001), while FVC z-score differences were not significant. Age, sex distribution, and smoking status were similar across groups. The distribution of primary respiratory diagnoses was broadly comparable. Collectively, severe pulmonary exacerbations manifested in individuals with greater clinical complexity, expressed as comorbidity burden and previous usage of healthcare resources, and worse airflow obstruction rather than differences in age, sex, smoking habits, or diagnostic labels.

**TABLE 1:** Main features of the study group: Comparisons between patients showing unplanned hospitalizations during the study period (n=50) and all the other patients (n=128)

| Variable | All other patients (n=128) | Patients with unplanned hospitalizations (n=50) | p-value |
| --- | --- | --- | --- |
| Age, Gender and Smoking habit |  |  |  |
| Age; mean (SD) | 66.96 (12.17) | 68.04 (12.01) | .596 |
| Male; n (%) | 50 (39.06) | 25 (50) | .237 |
| Female; n (%) | 78 (60.94) | 25 (50) |  |
| Current Smokers; n (%) | 16 (12.5) | 9 (18) | 0.304 |
| Former Smokers; n (%) | 73 (57.03) | 31 (62) |  |
| Never Smokers; n (%) | 39 (30.47) | 10 (20) |  |
| Main Respiratory Diagnosis, Co-morbidities and Forced Spirometry |  |  |  |
| COPD; n (%) | 60 (46.88) | 33 (66) | .166 |
| Asthma, n (%) | 11 (8.59) | 3 (6) |  |
| Bronchiectasis, n (%) | 19 (14.84) | 4 (8) |  |
| Severe asthma; n (%) | 38 (29.69) | 10 (20) |  |
| AMG score; mean (SD) | 71.09 (42.48) | 97.93 (50.96) | <.001 |
| AMG risk group - Very high risk; n (%) | 9 (7.03) | 11 (22) | .007 |
| AMG risk group – High risk; n (%) | 37 (28.91) | 19 (38) |  |
| AMG risk group – Moderate risk; n (%) | 54 (42.19) | 15 (30) |  |
| AMG risk group – Low risk; n (%) | 25 (19.53) | 3 (6) |  |
| AMG risk group – Very low risk; n (%) | 3 (2.34) | 1 (2) |  |
| FVC; z-score, mean (SD) | -1.09 (1.28) | -1.46 (1.34) | 0.121 |
| FEV1; z-score, mean (SD) | -1.85 (1.37) | -2.51 (1.42) | 0.009 |
| FEV1/FVC; mean (SD) | 62.63 (13.94) | 53.28 (17.16) | <.001 |
| Utilization of healthcare resources during the 12 months before retrospective clinical surveillance |  |  |  |
| All-cause hospitalizations; n (%) | 32 (25.00) | 31 (62.00) | <.001 |
| Unplanned hospitalizations; n (%) | 18 (14.06) | 29 (58.00) | <.001 |
| Total healthcare expenditure per patient in €; median (Q1-Q3) | 4581 (2,455 – 9,727) | 8,541 (5,157 – 14,057) | <.001 |

AMG score: Adjusted Morbidity Groups score, indicating co-morbidities and complexity; COPD: Chronic Obstructive Pulmonary Disease; FVC: Forced Vital Capacity; FEV1: Forced Expiratory Volume in 1 second; FEV1/FVC: Ratio of FEV1 to FVC; Total healthcare expenditure corresponds to total expenditure per year across all healthcare tiers.

### Household IAQ monitoring

#### Monitoring in practice: continuity and technical issues

Among the 183 active participants, 182 were successfully monitored over a 61-day period. However, four additional patients recorded fewer than 20 valid monitoring days and were therefore excluded from the analysis, resulting in the final analytic cohort of 178 participants (**Table 1**). Discontinuities in data monitoring (**Figure S6**) were mainly attributed to temporary device disconnections from the power supply, unstable Wi-Fi connectivity, and occasional sensor malfunctions or damage. Some incidents resolved spontaneously, while others required patient interaction following telephone assistance or on-site home visits. In a few cases, sensor replacement was needed to restore data transmission. On average, participants missed 5.0 (10.6) monitoring days, corresponding to 8.2% of all possible recording days.

#### Household air quality results and exposure patterns

**Table 2**summarizes the GO-AQS long-term thresholds for CO_2_, formaldehyde, and PM_2.5_, defining Low, Moderate, and Unhealthy exposure categories. It further reports, for each pollutant, the distribution of homes by category based on dwelling-level mean concentrations, and among these, the number of homes with any smoker present, and finally the overall monitoring time spent in each risk band. Among the 178 monitored homes, 92 (51.7%) exhibited at-risk pollution levels (moderate or unhealthy) for at least one monitored contaminant highlighting the prevalence of poor IAQ. CO_2_levels were generally within safe limits, with 163 (91.6%) homes classified as low risk and 14 (7.9%) falling into the moderate category. Only 1 (0.5%) home exceeded the unhealthy threshold for CO_2_. In contrast, formaldehyde pollution was more widespread, with 22 homes (12.4%) exceeding the low-risk threshold, 21 in the moderate category (11.9%) and 1 (0.5%) classified as unhealthy. PM_2.5_exposure was the most concerning, as 71 homes (40.1%) showed at-risk levels, with 58 in the moderate category (32.8%) and 13 in the unhealthy category (7.3%).

**TABLE 2:**
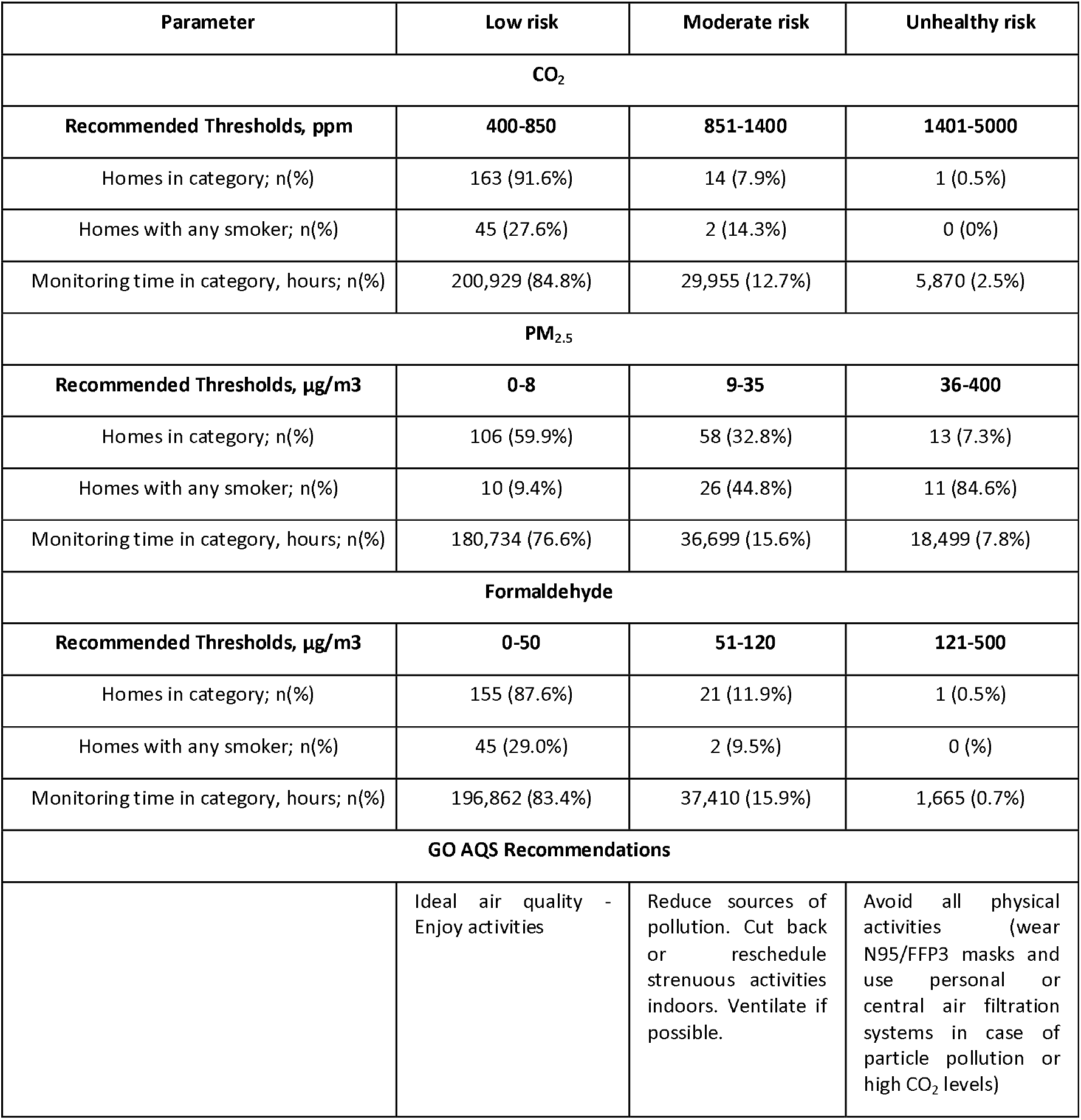
Results of household indoor air quality monitoring grouped by each of the three risk categories defined by the Global Open Air Quality Standards (GO AQS)^40^.

| Parameter | Low risk | Moderate risk | Unhealthy risk |
| --- | --- | --- | --- |
| **CO<sub>2</sub>** |  |  |  |
| **Recommended Thresholds, ppm** | **400-850** | **851-1400** | **1401-5000** |
| Homes in category; n(%) | 163 (91.6%) | 14 (7.9%) | 1 (0.5%) |
| Homes with any smoker; n(%) | 45 (27.6%) | 2 (14.3%) | 0 (0%) |
| Monitoring time in category, hours; n(%) | 200,929 (84.8%) | 29,955 (12.7%) | 5,870 (2.5%) |
| **PM<sub>2.5</sub>** |  |  |  |
| **Recommended Thresholds, µg/m3** | **0-8** | **9-35** | **36-400** |
| Homes in category; n(%) | 106 (59.9%) | 58 (32.8%) | 13 (7.3%) |
| Homes with any smoker; n(%) | 10 (9.4%) | 26 (44.8%) | 11 (84.6%) |
| Monitoring time in category, hours; n(%) | 180,734 (76.6%) | 36,699 (15.6%) | 18,499 (7.8%) |
| **Formaldehyde** |  |  |  |
| **Recommended Thresholds, µg/m3** | **0-50** | **51-120** | **121-500** |
| Homes in category; n(%) | 155 (87.6%) | 21 (11.9%) | 1 (0.5%) |
| Homes with any smoker; n(%) | 45 (29.0%) | 2 (9.5%) | 0 (%) |
| Monitoring time in category, hours; n(%) | 196,862 (83.4%) | 37,410 (15.9%) | 1,665 (0.7%) |
| **GO AQS Recommendations** |  |  |  |
|  | Ideal air quality -<br>Enjoy activities | Reduce sources of<br>pollution. Cut back<br>or reschedule<br>strenuous activities<br>indoors. Ventilate if<br>possible. | Avoid all physical<br>activities (wear<br>N95/FFP3 masks and<br>use personal or<br>central air filtration<br>systems in case of<br>particle pollution or<br>high CO<sub>2</sub> levels) |

As depicted in **Table 2**, smokers in dwellings varied across the different air quality risk categories, showing a clear association with PM_2.5_pollution levels, whereas smoker prevalence showed no increasing gradient across CO_2_or formaldehyde categories. In homes classified as low risk for PM_2.5_, only 10 (9.4%) had smokers, whereas this proportion increased to 26 (44.8%) in moderate-risk homes and reached 11 (84.6%) in unhealthy environments. This strong association (p <.001) suggests that smoking is a major contributor to fine particulate pollution indoors.

Analysing the time spent in each risk category suggests distinct emission patterns. PM_2.5_exposure showed a different pattern, with 59.9% of homes classified as low risk, but spending 76.6% of the time in this category (**Table 2**). This suggests acute pollution peaks driving PM_2.5_exposure. **Figure 2– Panel A**shows a representative 24-h PM_2.5_time series from an at-risk dwelling, illustrating typical indoor exposure dynamics. PM_2.5_pollution is manifested as pronounced diurnal peaks, exceeding “Moderate” or “Unhealthy” thresholds for short periods, potentially coinciding with activities such as cooking, cleaning (particularly sweeping or dusting, which resuspends particles), and, most prominently, tobacco use indoors. Other episodic sources included the burning of incense or candles. Nighttime concentrations tended to decrease sharply, consistent with particle sedimentation in the absence of human activity.

**Figure 2:**
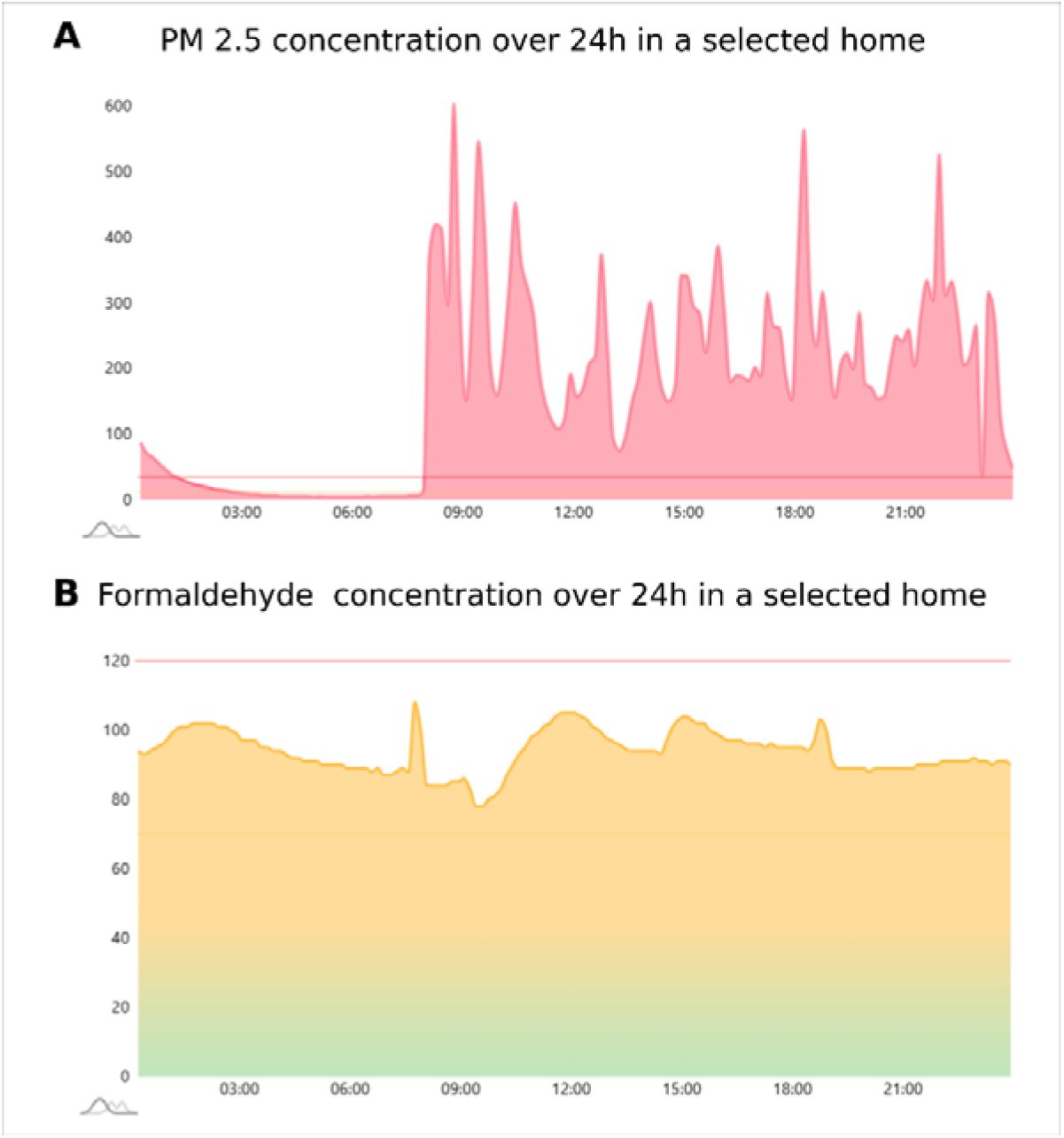
Twenty-four-hour indoor concentration profiles in selected at-risk homes (μg/m^3^). Panel A) PM2.5 exposure. Panel B) Formaldehyde exposure. The exposure was recorded at 10-min intervals with MICA-IBIOT LCS; screenshots exported from the myInbiot monitoring platform.

**Figure 2– Panel B**presents a 24-h monitoring segment from a dwelling representative of high formaldehyde contamination. Conversely, Formaldehyde exposure appears to be continuous, with 87.6% of homes classified as low risk, while spending only 83.4% of the monitored time in this category (**Table 2**). This behaviour reflects passive, continuous emissions, off-gassing, from materials such as furniture, construction products, and household goods, rather than direct links to occupant behaviours. These emissions were not associated with short-term activity peaks but could be influenced by environmental conditions such as temperature and ventilation.

As shown in **Figure 3**, the distributions of IAQ risk categories for CO_2_, PM_2.5_and formaldehyde were similar in patients that suffered severe pulmonary exacerbations requiring hospitalisation during the previous year of IAQ assessment and those without events. Only, PM_2.5_showed a modest, non-statistically significant, shift toward higher PM_2.5,_exposure among hospitalised patients. Similar distributions were observed when stratifying by all-cause emergency department visits.

**Figure 3:**
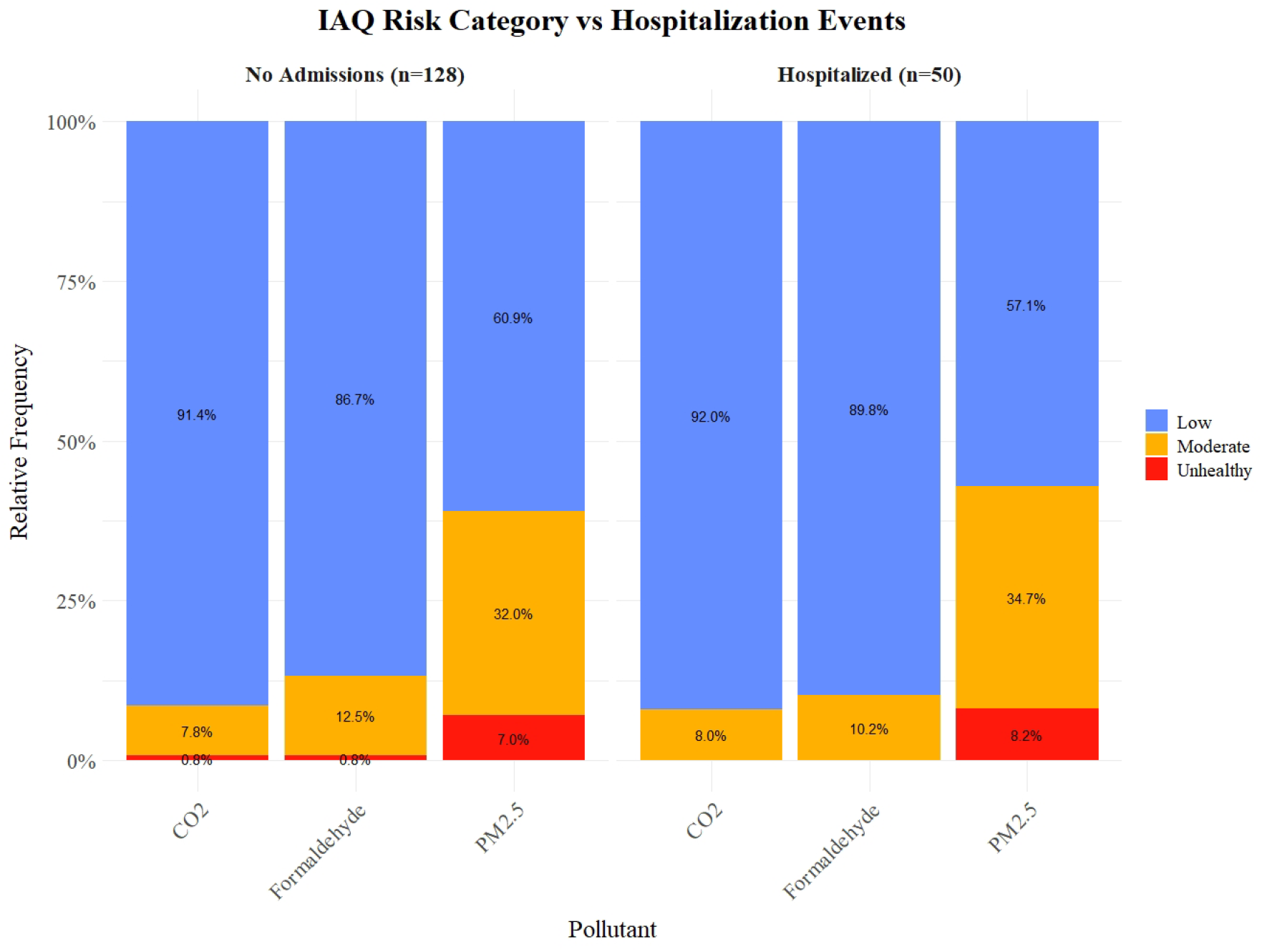
IAQ risk categories by pollutant and hospitalisation status. Stacked bars show the proportion of dwellings in Low, Moderate, and Unhealthy GO-AQS categories for each pollutant (CO_2_, formaldehyde, PM25), stratified by patients with no admissions (n=128) versus those hospitalised for severe pulmonary exacerbations (n=50). Percentages within bars indicate the share of homes per risk category in each subgroup.

## Discussion

This study reports the operational evaluation of LCS for home-based IAQ assessment and explores clinical applicability of this type of sensors in patients with chronic respiratory disease. This research contributed specifically to two areas: 1) Feasibility and readiness of LCS-supported household IAQ monitoring; 2) the role of IAQ in preventive and personalized care.

### Operational feasibility and readiness of LCS

#### Technological readiness

In field deployment, the LCS units provided stable, approximately linear responses within the relevant concentration ranges for particulate matter and formaldehyde, and consistently captured relative changes and temporal patterns, enabling reliable characterisation of household exposure profiles. These observations are consistent with independent evaluations of LCS performance^20–22^. In contrast, total VOC readings tended to overestimate concentrations and showed cross-interference, limiting their suitability for absolute quantification. Nevertheless, they retained qualitative utility in identifying high-emission events (see **Supplementary Material**for details on laboratory assessment of the total VOC sensor). CO_2_levels, on the other hand, primarily reflected indoor occupancy and ventilation status rather than pollution per se, and are best interpreted as indicators of air stagnation or insufficient ventilation within the home.

#### Usability considerations

Some participants experienced temporary data losses due to device disconnections, Wi-Fi instability, sensor malfunction, or prolonged absences from home during holiday periods. While most incidents were minor and easily resolved, either automatically, through patient support, or by on-site visits, these interruptions highlight the operational demands of maintaining continuous long-term monitoring. Therefore, from an operational perspective, continuous long-term household IAQ monitoring is not cost-effective. Considering the maintenance and synchronization requirements of IAQ monitors, together with the relative stability of exposure patterns once pollution sources are identified, indefinite monitoring offers limited additional value. We therefore recommend time-limited IAQ assessments, either as an initial screening for patients with difficult-to-control COPD or asthma, or as a follow-up to evaluate the impact of targeted environmental interventions.

#### Future improvements

Clinical decision-making would be strengthened by multi-pollutant sensing that includes oxidants (NOx, O_3_). Oxidant gases are associated with respiratory morbidity and can amplify PM-related oxidative stress and airway inflammation when present together, potentially increasing the risk of exacerbations^47^; a multi-pollutant view is therefore biologically and clinically coherent. Incorporating validated NO_2_/O_3_sensing could improve early detection of hazardous indoor conditions, trigger timely prevention of acute exacerbations. Equally important is the refinement of VOC sensing. Current VOC sensors are qualitative and non-specific, responding to a wide range of compounds with overlapping signal patterns that make it difficult to identify individual sources or quantify absolute concentrations. This lack of specificity limits their interpretability for clinical or epidemiological purposes.

### The role of IAQ in preventive and personalized care

#### Prevalence and composition of household air pollution

Impaired household IAQ is highly prevalent. More than half of the monitored dwellings exhibited average concentrations above recommended thresholds for at least one of the monitored pollutants. Fine particulate matter was the dominant contributor to poor IAQ, and we identified indoor smoking as a key driver of particulate pollution. Nevertheless, elevated PM_2.5_in non-smoker dwellings indicates additional relevant sources, most notably cooking emissions, combustion appliances, and cleaning/resuspension, underscoring the need for context-specific mitigation. These results emphasise the relevance of PM exposure beyond tobacco smoke and support tailored recommendations to reduce particulate burden, that primarily rely on behavioural change, as most relevant sources are linked to everyday human activities. In contrast, formaldehyde contamination followed a distinct pattern characterised by persistent, thermally dependent off-gassing from structural and furnishing materials. Because such emissions are continuous and largely independent of occupant behaviour, mitigation should prioritise source control using low-emission materials, sustained ventilation, and temperature regulation to limit volatilisation. In severe cases, gas-phase removal is feasible with sorbent media (e.g., activated carbon/chemisorption cartridges).

#### Environmental and clinical determinants of respiratory exacerbations

Hospitalization risk among complex chronic respiratory patients is primarily determined by the cumulative burden of multimorbidity, the severity of pulmonary impairment, and the individual history of exacerbations^48,49^, and environmental exposures seem to play a secondary role relative to intrinsic clinical determinants in populations characterized by advanced disease stages. However, the absence of an immediate relationship between short-term IAQ and severe exacerbations should not be interpreted as a lack of relevance of indoor exposures. Rather, it reflects the temporal divide between environmental exposure and clinical manifestation, as well as the multifactorial nature of exacerbation risk. In this regard, it is well acknowledged that chronic diseases evolve under the cumulative influence of the exposome, the totality of environmental and behavioural exposures experienced over time, acting as long-term modifier of disease trajectory^50^. In addition, the intrinsic characteristics of the study design may have limited the ability to detect measurable associations at this stage. By focusing exclusively on hospitalisations for severe exacerbations, the analysis intentionally captured only the most critical outcomes, thereby excluding a large proportion of mild and moderate events typically managed in community or home settings. Moreover, the study did not account for infectious triggers, recognised as major drivers of acute respiratory deterioration, or for oxidant pollutants that may act synergistically with particulate matter, chemical compounds, and bioaerosols to amplify airway inflammation and exacerbate symptoms^51^.

#### Towards Hybrid Care: Digital and clinical strategies for prevention

Preventing hospitalisations in patients with advanced chronic respiratory disease requires a shift toward early detection and proactive management of community-based exacerbations. This approach is at the core of the ongoing hybrid care intervention implemented in the study cohort ^28^. That combines remote and in-person care, coordinated by a nurse case manager and enabled by a digital adaptive case management platform^52^. In daily clinical practice, this structure has the potential to enhance the early recognition and management of acute episodes in the community, thereby reducing hospital dependence and enabling more continuous, patient-centred follow-up. While these capabilities remain in the process of clinical validation, preliminary experience suggests that integrating routine assessments with remotely collected data on lung function, symptom trajectories, and heart rate variability (HRV) can provide a more objective basis for identifying early signs of deterioration. The evolving profiles of these parameters from baseline to recovery may, in the future, support data-driven stratification of exacerbation risk and open new avenues for personalised, anticipatory management within hybrid care models.

### Strengths, limitations and future research

This study represents one of the first large-scale deployments of LCS for household IAQ monitoring in patients with advanced chronic respiratory disease, integrated within a digitally enabled hybrid care model. The results provide early but solid evidence on the feasibility, robustness, and clinical applicability of remote IAQ assessment, offering practical insights that can guide both research and clinical adoption.

Building on these findings, the ongoing work will expand toward a prospective, longitudinal analysis that integrates environmental and clinical dimensions across the full follow-up period, incorporates broader health outcomes and community-level impact indicators, and refines exacerbation evaluation through patient-reported outcomes and physiological metrics such as Oscillometry and heart rate variability (HRV). Parallel efforts will focus on characterising dwelling structures and pollution sources following IPCHEM guidelines, and on translating monitoring into action through targeted patient-centred initiatives, including educational and tobacco-cessation campaigns, real-time feedback tools to promote ventilation awareness, and a pilot household air-filtration protocol in highly polluted homes. Together, these next steps aim to consolidate the clinical value of IAQ monitoring and support its integration into preventive, digitally enabled respiratory care.

## Conclusions

This study demonstrates the feasibility, reliability, and clinical relevance of using LCS for short-term household IAQ screening in patients with chronic respiratory disease. Household air pollution was found to be highly prevalent, predominantly driven by fine PM and indoor smoking. These findings highlight the need to address indoor pollution as a modifiable exposure and open new clinical perspectives for integrating environmental assessment into preventive respiratory care.

## Abbreviations

ACM: *Adaptive Case Management*
AISBE: *Àrea Integral de Salut Barcelona-Esquerra*
AMG: *Adjusted Morbidity Groups*
CH_2_O: *Formaldehyde*
CHSS: *Catalan Health Surveillance System*
CO_2_: *Carbon dioxide*
COPD: *Chronic Obstructive Pulmonary Disease*
ED: *Emergency Department*
FEV_1_: *Forced Expiratory Volume in one second*
FVC: *Forced Vital Capacity*
GO-AQS: *Global Open Air Quality Standards*
HCB: *Hospital Clínic de Barcelona*
HRV: *Heart Rate Variability*
IAQ: *Indoor Air Quality*
ICD-10-CM: *International Classification of Diseases, 10th Revision, Clinical Modification*
LCS: *Low-Cost Sensors*
NOlll: *Nitrogen Oxides*
O_3_: *Ozone*
OAQ: *Outdoor Air Quality*
PM: *Particulate Matter*
PDSA: *Plan–Do–Study–Act*
QoL: *Quality of Life*
SD: *Standard Deviation*
STROBE: *Strengthening the Reporting of Observational Studies in Epidemiology*
VOCs: *Volatile Organic Compounds*

## Data Availability

The datasets generated and/or analysed during the current study contain sensitive patient information and will not be openly distributed. However, anonymized data may be made available upon reasonable request to the corresponding author, subject to institutional data sharing agreements and ethical approval.

## Authors’ Contributions

RGC, JJR, IC, JF, and EA conceived and supervised the study. RGC developed the statistical analysis plan, performed the data analysis, and created the figures. AGL was responsible for patient recruitment, cohort follow-up, and data collection as part of the study’s nursing role. NS, NSR, MS and EA represented the medical teams from both hospital and primary care, contributing to the study’s clinical aspects. MF provided expertise as a representative of the monitoring technology providers. AA, JF, and AR contributed their expertise in IAQ and performed the laboratory validation of the environmental sensors. RF served as an advisor on Oscillometry procedures. EV and JPJ facilitated access to regional health data and provided statistical support. RGC, AGL, JR, IC, and EA led the manuscript drafting. All authors contributed to the writing, reviewed the manuscript and approved the final version.

## Acknowledgements

The K-HEALTHinAIR project funded this study, Grant Agreement nº 101057693, under a European Union’s Call on Environment and Health (HORIZON-HLTH-2021-ENVHLTH-02).

This research was also supported by the Catalan Government and the Catalan Department of Research and Universities under contract 2021 SGR 00326.

## Disclaimer

Views and opinions expressed are, however, those of the authors only and do not necessarily reflect those of the European Union or the European Health and Digital Executive Agency as granting authority. Neither the European Union nor the granting authority can be held responsible.

## Patient and Public Involvement

Patients or the public were not involved in the design, or reporting, or dissemination plans of our research.

## Competing Interests

IC and JR hold shares in Health Circuit SL. JR contributes to the Astra-Zeneca Global Oscillometry Advisory Board. All other authors declare no conflicts of interest.

MF is the Chief Scientific Officer of inBioT, the manufacturer of the MICA sensors utilized in this study.

