## Supplementary Material for "Should Household Air Quality Monitoring Be Considered in Selected Asthma & COPD Patients?"

### Household Indoor Air Quality (IAQ) monitoring with Low-Cost Sensors (LCS)

#### 1.1. Background and Objectives

Low-cost sensors (LCS) have become a viable and increasingly popular solution for monitoring IAQ. Their low cost, scalability, and ease of installation make them particularly suitable for longitudinal studies and spatially distributed IAQ assessments. Nonetheless, these benefits are tempered by recognized technical limitations that compromise their precision and comparability to reference-grade instruments. Sensor performance can vary markedly depending on the pollutant measured, the underlying sensing technology, and the environmental conditions in which they operate. These factors must be carefully considered when interpreting results and conducting exposure-response analyses. We assessed the accuracy and reproducibility of LCS for detecting particulate matter (PM) at various sizes (1/2.5/10), formaldehyde (CH₂O), and total volatile organic compounds (tVOCs).

#### 1.2 Results of the LCS assessment

###### 1.2.1 Technical specifications of integrated environmental sensors

The environmental monitoring system used in this study was the MICA Plus (inBiot), equipped with a comprehensive suite of sensors for assessing IAQ. **Table S1** summarizes the specifications of the sensors incorporated into the MICA device, including their type, measuring ranges, resolution, and accuracy provided by the manufacturer.

**Table S1. Measurement range, resolution, accuracy, and sensing technologies of the sensors integrated into the MICA-inBiot indoor air quality monitoring devices.**

| **Parameter** | **Technology** | **Measurement Range** | **Resolution** | **Accuracy** |
| --- | --- | --- | --- | --- |
| **Temperature** | Silicon bandgap | -40 to 125 °C | 0.1 °C | ± 0.5 °C |
| **Relative Humidity (RH)** | Capacitive | 0 to 100 % RH | 1 % RH | ± 2 % RH |
| **CO₂** | NDIR (non-dispersive infrared) | 400 to 10,000 ppm | 1 ppm | ± (30 ppm + 3 % of measured value) |
| **TVOC** | MOx (metal-oxide) | 0 to 2383 ppb | 1 ppb | ± 15 % of measured value |
| **Particulate Matter** | Optical laser | 0 to 1000 μg/m³ | 1 μg/m³ | **PM₁ / PM₂.₅**: ± (5 μg/m³ + 5 % mv) (0–100 μg/m³), ± 10 % mv (101–1000 μg/m³)  **PM₄ / PM₁₀**: ± 25 μg/m³ (0–100 μg/m³), ± 25 % mv (101–1000 μg/m³) |
| **Formaldehyde** | Electrochemical | 0 to 1000 ppb | 1 ppb | ± 20 ppb or ± 20 % mv (whichever is greater) |

Further details about the long-term performance and environmental robustness of the integrated sensors are provided in **Table S2**. This includes information on expected lifespan, annual drift, and the operational temperature and humidity ranges for each sensor type.

****

**Table S2. Sensor lifespan, drift, and environmental operating ranges for MICA-inBiot indoor air quality monitoring devices.**

| **Parameter** | **Lifespan** | **Drift** | **Operating Temperature Range** | **Operating Humidity Range** |
| --- | --- | --- | --- | --- |
| **Temperature** | — | < 0.03 °C/year | -40 to 105 °C | 0 to 100 % RH |
| **Relative Humidity** | — | < 0.2 % RH/year | -40 to 105 °C | 0 to 100 % RH |
| **CO₂** | > 10 years | — | -10 to 50 °C | 0 to 95 % RH |
| **TVOC** | > 10 years | — | -10 to 50 °C | 0 to 95 % RH |
| **Particulate Matter (PM)** | > 10 years | 0–100 μg/m³: ± 1.25 μg/m³/year  100–1000 μg/m³: ± 1.25 % mv/year | -10 to 50 °C | 0 to 95 % RH |
| **Formaldehyde** | > 6 years | < 5 ppb or < 5 % of measured value per year (whichever is greater) | 0 to 50 °C | 10 to 95 % RH |

##### 1.2.2 PM 1/2.5/10

The MICA-inBiot devices used in this study incorporate the Sensirion SPS30 optical particle counter (<https://sensirion.com/products/catalog/SPS30>). This sensor estimates PM concentrations by measuring light scattered by airborne particles passing through a laser beam. The intensity and angle of scattered light are interpreted via proprietary algorithms, converting size-classified particle counts into mass concentrations (µg/m³) for PM1, PM2.5, and PM10, based on assumed particle density and shape.

*Experimental validation:*

Previous studies conducted by Aguado et al. ^1^ focused on the evaluation of the performance of the commercial sensor SPS30 to measure PM (PM 1.0, PM2.5 and PM10.0) in laboratory conditions by comparing the measurements against calibrated reference equipment.

The SPS30 sensor complied with the manufacturer's stated performance criteria for PM1. However, a systematic underestimation was observed for both PM2.5 and PM10 across trials. Notably, strong linear correlations were maintained between the low-cost sensor and the reference instrument (R² = 0.87 for PM10; up to R² = 0.99 for PM1), indicating preserved trend fidelity despite bias in absolute values. Additionally, the temporal stability of sensor response was assessed over 6000 hours of continuous operation, with no evident signs of drift, supporting the device’s reliability for long-term monitoring.

A further experimental validation campaign was conducted to evaluate the accuracy, linearity, and inter-sensor consistency of the SPS30 sensor under non-controlled indoor conditions. Environmental parameters were not artificially stabilized, allowing the evaluation of sensor behavior across naturally varying conditions. Six devices were deployed simultaneously to evaluate intra-model reproducibility, while two calibrated comparison-grade monitors served as reference devices for assessing accuracy and trend agreement. All devices operated concurrently in the same microenvironment with synchronized data collection intervals.

Under these indoor environmental conditions, SPS30-based sensors demonstrated high linearity and low inter-unit variability (R^2^=0.77) for PM1, indicating reliable intra-model consistency for fine particle monitoring (**Figure S1**). However, the sensors tended to underestimate particle concentrations compared to the reference values, particularly at higher PM1 levels. For PM2.5, although a systematic underestimation was observed, particularly above 10 µg/m³, the readings of the multiple sensors remained strongly correlated (R^2^=0.78) (**Figure S2**), supporting their use for trend analysis with potential need for empirical correction factors for accurate exposure analysis. By contrast, while PM10 estimates remain correlated (R^2^=0.87) among them, they showed increasing divergence from the reference at higher concentrations, with evidence of non-linear bias and greater variability across sensor units (**Figure S3**). This divergence is consistent with the sensor's known limitations in detecting coarse particles, as PM10 estimates are extrapolated from smaller particle counts and not directly measured.

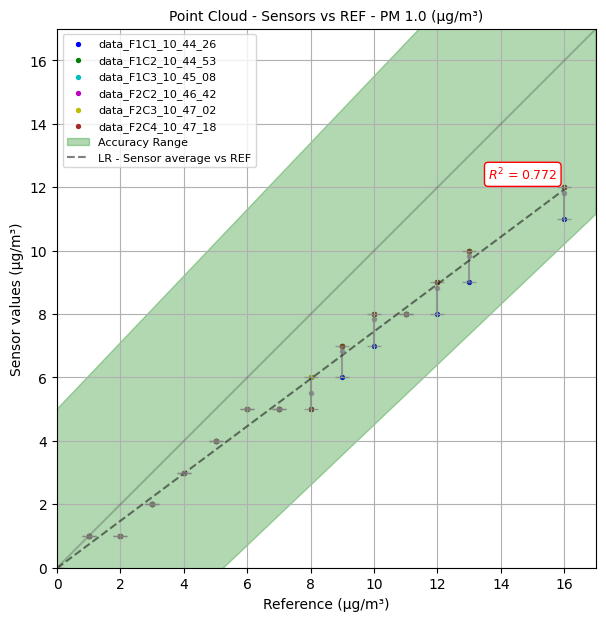

***Figure S1. Experimental validation of particulate matter (PM1) sensors (SPS30) integrated in MICA devices against comparison-grade monitors serving as reference devices.*** *Scatter plot comparing the response of six SPS30-based MICA sensors (y-axis) to PM1 reference measurements (x-axis). Each point reflects the mean reading of an individual device at a given reference level, with vertical bars representing inter-sensor variation, color-coded per device ID. The shaded area indicates the manufacturer’s specified precision bounds (±5 µg/m³ or ±10% m.v., depending on concentration), based on factory calibration under standard conditions*

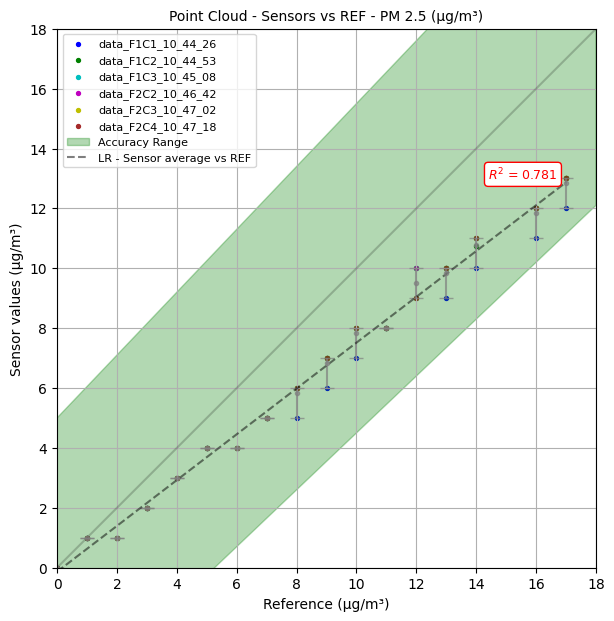

***Figure S2. Experimental validation of particulate matter (PM2.5) sensors (SPS30) integrated in MICA devices against comparison-grade monitors serving as reference devices.*** *Scatter plot comparing the response of six SPS30-based MICA sensors (y-axis) to PM2.5 reference measurements (x-axis). Each point reflects the mean reading of an individual device at a given reference level, with vertical bars representing inter-sensor variation, color-coded per device ID. The shaded green area delineates the manufacturer’s nominal accuracy range (±5 µg/m³).*

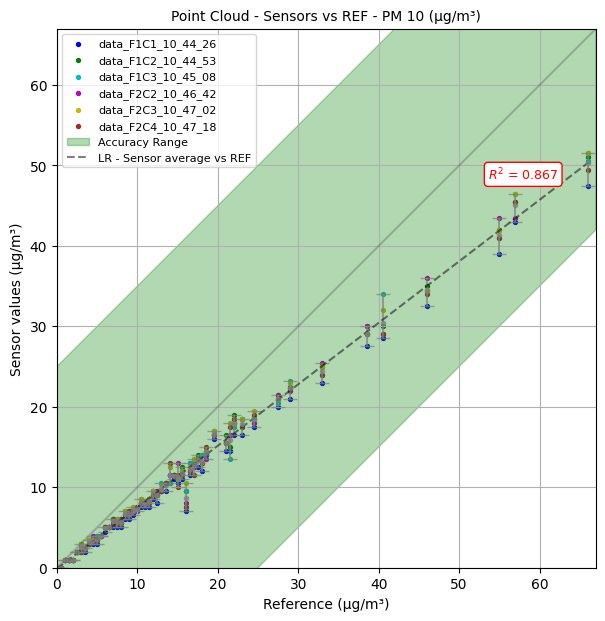

***Figure S3. Experimental validation of particulate matter (PM10 estimates) sensors (SPS30) integrated in MICA devices against comparison-grade monitors serving as reference devices.*** *Scatter plot comparing the response of six SPS30-based MICA sensors (y-axis) to PM10 reference measurements (x-axis). Each point reflects the mean reading of an individual device at a given reference level, with vertical bars representing inter-sensor variation, color-coded per device ID. The shaded area indicates the manufacturer’s specified precision bounds (±5 µg/m³ or ±10% m.v., depending on concentration), based on factory calibration under standard conditions*

###### 1.2.3 Formaldehyde

The MICA-inBiot device integrates the compact electrochemical sensor Sensirion SFA30 (<https://sensirion.com/products/catalog/SFA30>), designed for continuous monitoring of airborne formaldehyde. This sensor operates by generating an electric current proportional to the concentration of formaldehyde through redox reactions at the electrode surface.

*Experimental validation:*

Previous validation studies have shown that the SFA30 sensor performs adequately under baseline indoor conditions, with acceptable agreement to reference devices despite some initial variability⁵. While offering good selectivity and response time, key limitations include sensor drift, sensitivity to environmental conditions, and cross-sensitivity to other gases (e.g., CO, NOₓ, VOCs)^2^. These effects may be exacerbated under low-concentration conditions, where signal-to-noise ratio decreases, potentially compromising accuracy^2,3^. Nonetheless, literature supports the use of electrochemical formaldehyde sensors like the SFA30 for screening significant deviations and detecting exposure events, particularly when used with proper calibration strategies and environmental controls ^2,4^.

Within this study, to assess the accuracy and selectivity of the Sensirion SFA30 electrochemical formaldehyde sensor integrated in the MICA-inBiot IAQ monitors, a series of controlled exposure tests were conducted in a 1 m³ environmental chamber. The concentration ranges evaluated in the chamber (FA ≈ 55–78 µg/m³; AC ≈ 24–45 µg/m³) are relevant for real-world indoor environments, especially in healthcare or occupational settings. Each trial involved the injection of known concentrations of formaldehyde, accompanied by residual levels of acetaldehyde, which could not be fully eliminated despite air purification. Both formaldehyde and acetaldehyde were simultaneously sampled and quantified using a validated reference method based on DNPH (2,4-dinitrophenylhydrazine) cartridge collection followed by HPLC analysis.

As summarized in **Table S3**, the sensor readings generally overestimated formaldehyde concentrations when compared to formaldehyde-only reference values. However, these deviations were substantially reduced when the combined formaldehyde and acetaldehyde concentrations were considered, indicating a detectable cross-sensitivity to acetaldehyde, consistent with the behavior observed in other electrochemical sensors. While the sensor variability across devices slightly exceeded the manufacturer’s specified accuracy threshold in some instances, the overall pattern of response remained coherent and concentration-dependent, with sensor readings showing stable trends across increasing exposure levels.

**Table S3. Summary of formaldehyde concentrations measured by twelve SFA30 sensors (integrated into MICA Plus IAQ monitors) under controlled laboratory conditions.**

| **Trial** | **SFA30 Sensor (µg/m³); mean (SD)** | **Formaldehyde Reference (µg/m³)** | **Formaldehyde + Acetaldehyde Reference (µg/m³)** | **Δ vs Formaldehyde; Abs. (Rel. %)** | **Δ vs Formaldehyde + Acetaldehyde; Abs. (Rel. %)** |
| --- | --- | --- | --- | --- | --- |
| **T1** | 84.08 (16.33) | 56 | 79 | 28.08 (150.15%) | 5.08 (106.43%) |
| **T2** | 83.17 (16.39) | 57 | 93 | 26.17 (145.91%) | -9.83 (89.43%) |
| **T3** | 122.33 (25.95) | 78 | 123 | 44.33 (156.84%) | -0.67 (99.46%) |
| **T4** | 121.5 (27.53) | 76 | 119 | 45.5 (159.87%) | 2.5 (102.1%) |

*Four exposure scenarios (T1–T4) with different formaldehyde and acetaldehyde concentrations were tested. For each trial, the table reports the mean and standard deviation (SD) across all 12 sensors, alongside reference values for FA alone and for combined FA+AC (measured via DNPH/HPLC). Absolute and relative deviations from both references are included to assess accuracy and potential cross-sensitivity effects.*

A further experimental validation campaign was conducted to evaluate the accuracy, linearity, and inter-sensor consistency of the SFA30 sensor under non-controlled indoor conditions. As shown in **Figure S4,** the formaldehyde sensor response demonstrated good tracking of increasing reference concentrations (R^2^ = 0.83), but with a consistent positive bias across all exposure levels. Most sensor readings fell within the manufacturer-defined accuracy range; however, they tended to overestimate formaldehyde concentrations, especially at higher exposures. Importantly, inter-sensor variability was evident, particularly at mid-to-high concentration levels (10–30 µg/m³), as reflected by the visible spread between individual sensor curves and the magnitude of standard deviation bars. While the magnitude of the bias varied across devices, all sensors exhibited the same directional trend, indicating preserved monotonicity and consistent relative ranking of exposure levels.

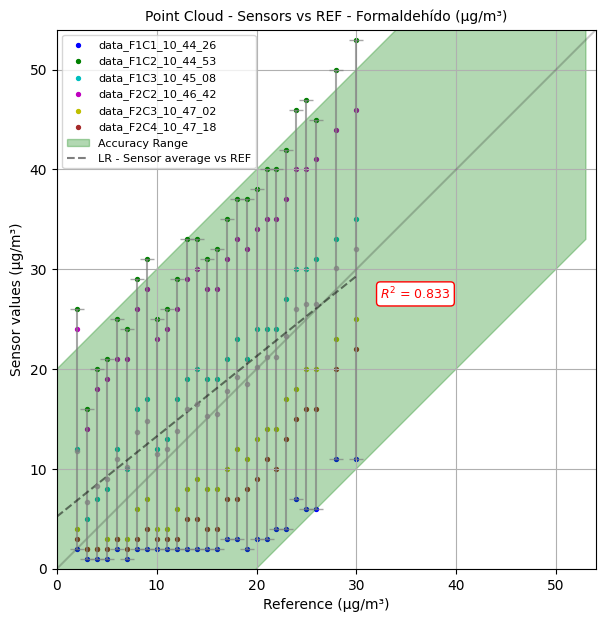

***Figure S4. Experimental validation of formaldehyde sensors (SFA30) integrated in MICA devices against comparison-grade monitors serving as reference devices.*** *Scatter plot comparing the response of six SFA30-based MICA sensors (y-axis) to formaldehyde reference measurements (x-axis). Each point reflects the mean reading of an individual device at a given reference level, with vertical bars representing inter-sensor variation, color-coded per device ID. The shaded green area delineates the manufacturer’s nominal accuracy range (±25 µg/m³).*

###### 1.2.3 Total Volatile Organic Compounds (tVOCs)

The MICA-inBiot devices used in this study incorporate Sensirion SGP40 metal-oxide (MOX) sensors (<https://sensirion.com/products/catalog/SGP40>) for continuous monitoring of tVOCs. These low-cost sensors offer a general indication of indoor air pollution by responding to a broad spectrum of oxidizable gases, including various VOCs and, to a lesser extent, some inorganic compounds. However, they lack chemical specificity and cannot differentiate between individual substances present in the air.

The sensor output consists of a composite index generated from signal fluctuations processed through proprietary algorithms calibrated by the manufacturer. As a result, the output is not directly linked to absolute concentration values or established health-based thresholds, limiting its interpretability. Furthermore, the sensor's performance is highly influenced by environmental conditions such as temperature and relative humidity, and it requires conditioning and baseline adaptation to achieve a stable response under varying ambient conditions.

Although Sensirion SGP40 provides a VOC Index and supports calibration routines, the data produced remains qualitative in nature. Given the sensor’s inherent non-linearity, cross-sensitivity, susceptibility to environmental interference, and lack of standardization, the study did not report tVOC data in the final analysis. This decision reflects the need for more robust, selective, and quantitatively reliable sensing technologies when compound-specific information or accurate exposure assessments are required.

###### 1.2.4 Key findings and recommendations

**Particulate Matter (PM1, PM2.5, PM10):**

1. **PM1:** Excellent agreement with reference measurements under laboratory conditions, showing high linearity and minimal inter-sensor variability. Suitable for both trend analysis and approximate quantification in typical indoor settings.
2. **PM2.5:** A systematic underestimation was observed, particularly above 10 µg/m³, but linear trends were preserved. Performance is adequate for trend tracking, though empirical correction factor may be required for precise exposure quantification in homes.
3. **PM10:** Marked divergence at concentrations above 40 µg/m³, with increasing non-linearity and inter-sensor variability. Indicative use is possible, but precision is limited without calibration.

**Formaldehyde:**

1. The SFA30 sensor showed consistent overestimation relative to formaldehyde-only reference data but improved agreement when considering combined formaldehyde and acetaldehyde, indicating a detectable cross-sensitivity. Both are well recognized carcinogen compounds.
2. Despite inter-sensor variability and limited absolute accuracy (especially at low concentrations), the device demonstrated stable monotonic response and preserved ranking across exposure levels.
3. Results confirmed its suitability as a screening tool for identifying potential IAQ concerns, particularly when formaldehyde levels approach or exceed 50 µg/m³.

**Total Volatile Organic Compounds (tVOCs):**

The MOX sensors used for tVOC monitoring lack specificity and suffer from drift, cross-sensitivity, and environmental sensitivity. Due to these limitations, tVOC data were not included in the present analysis.

**Overall:**

Despite the limitations, LCS remain highly valuable tools for environmental health monitoring. While they may not always provide perfectly accurate absolute concentrations, their response is typically linear and stable enough to reliably detect relative changes and pollution trends over time. This makes LCS particularly well-suited for applications such as identifying hot-spots, monitoring communities, and assessing temporal variability in IAQ. When used as indicative monitors, LCS can effectively raise awareness and support screening-level assessments. They might be powerful enablers of risk stratification and prioritization, especially when integrated into broader health strategies in vulnerable populations, such as complex respiratory patients. Although calibration and uncertainty analysis are necessary to translate their outputs into precise concentrations, their screening capabilities alone are often sufficient to trigger appropriate mitigation measures.

**Action plan in polluted homes:**

**Figure S5** outlines specific intervention strategies based on the primary pollutant source: particulate pollution or formaldehyde pollution. For particulate pollution, interventions include identifying and eliminating indoor smoking, promoting smoking cessation, and improving air quality in smoke-free homes through source identification, source removal, and the use of HEPA filtration systems. For formaldehyde pollution, the strategy involves source identification, removal, and the use of air purifiers with activated carbon. All identified at-risk homes will undergo validation of measurements to confirm pollutant levels. Additionally, residents of homes classified as low risk will receive indoor air quality education through digital tools to promote continued healthy practices.

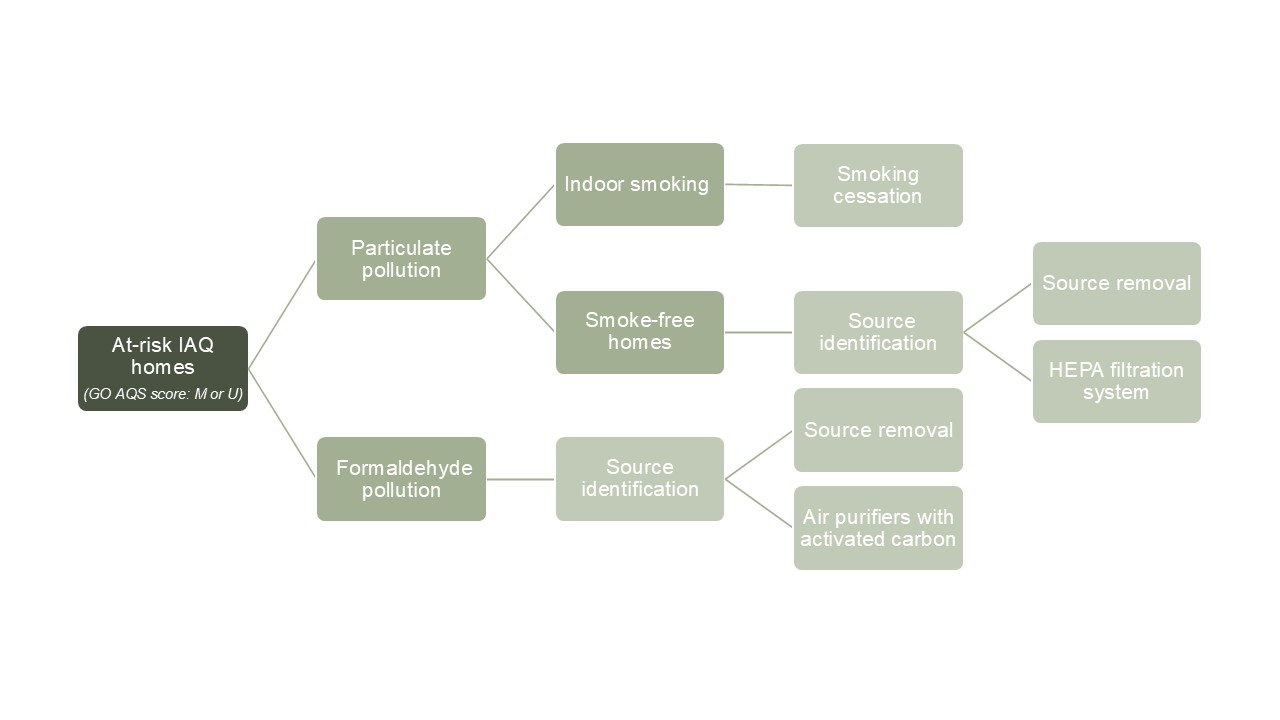
***Figure S5. Targeted action plan for homes identified with consistently poor indoor air quality (IAQ) based on the GO AQS scoring system, categorized as Moderate (M) or Unhealthy (U).*** *The plan outlines specific intervention strategies based on the primary pollutant source: particulate matter or formaldehyde pollution.*

###### 1.2.5 Monitoring adherence and data continuity

**Figure S6** illustrates the continuity of household air-quality monitoring between April 1 and May 31, 2025. During this 61-day follow-up, 182 participants were monitored, with an average of 5.0 (10.6) missing monitoring days per participant. As shown in the figure, 124 participants (68.1%) maintained full data continuity without any missing days, 41 (22.5%) missed between 1 and 10 days, and 17 (9.3%) experienced longer data gaps exceeding 10 days.

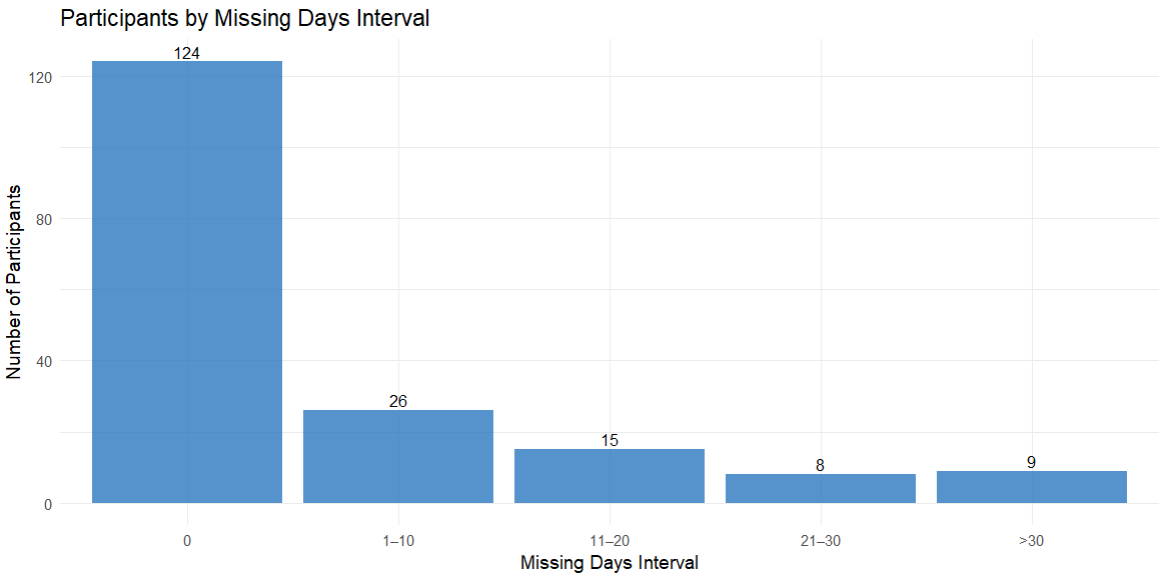

***Figure S6. Distribution of participants according to the number of missing monitoring days during the follow-up period (April 1 – May 31, 2025).***
